# Improving the Prediction of Death from Cardiovascular Causes with Multiple Risk Markers

**DOI:** 10.1101/2023.01.21.23284863

**Authors:** Xin Wang, Kelly M. Bakulski, Samuel Fansler, Bhramar Mukherjee, Sung Kyun Park

**Author notes:** **Correspondence to Xin Wang,** Department of Epidemiology, University of Michigan, 6630 SPH I, 1415 Washington Heights, Ann Arbor, Michigan 48109-2029.

## Abstract

**Background:** Traditional risk factors including demographics, blood pressure, cholesterol, and diabetes status are successfully able to predict a proportion of cardiovascular disease (CVD) events. Whether including additional routinely measured factors improves CVD prediction is unclear. To determine whether a comprehensive risk factor list, including clinical blood measures, blood counts, anthropometric measures, and lifestyle factors, improves prediction of CVD deaths beyond traditional factors.

**Methods:** The analysis comprised of 21,982 participants aged 40 years and older (mean age=59.4 years at baseline) from the National Health and Nutrition Examination Survey (NHANES) from 2001 to 2016 survey cycles. Data were linked with the National Death Index mortality data through 2019 and split into 80:20 training and testing sets. Relative to the traditional risk factors (age, sex, race/ethnicity, smoking status, systolic blood pressure, total and high-density lipoprotein cholesterol, antihypertensive medications, and diabetes), we compared models with an additional 22 clinical blood biomarkers, 20 complete blood counts, 7 anthropometric measures, 51 dietary factors, 13 cardiovascular health-related questions, and all 113 predictors together. To build prediction models for CVD mortality, we performed Cox proportional hazards regression, elastic-net (ENET) penalized Cox regression, and random survival forest, and compared classification using C-index and net reclassification improvement.

**Results:** During follow-up (median, 9.3 years), 3,075 participants died; 30.9% (1,372/3,075) deaths were from cardiovascular causes. In Cox proportional hazards models with traditional risk factors (C-index=0.850), CVD mortality classification improved with incorporation of clinical blood biomarkers (C-index=0.867), blood counts (C-index=0.861), and all predictors (C-index=0.871). Net CVD mortality reclassification improved 13.2% by adding clinical blood biomarkers and 12.2% by adding all predictors. Results for ENET-penalized Cox regression and random survival forest were similar. No improvement was observed in separate models for anthropometric measures, dietary nutrient intake, or cardiovascular health-related questions.

**Conclusions:** The addition of clinical blood biomarkers and blood counts substantially improves CVD mortality prediction, beyond traditional risk factors. These biomarkers may serve as an important clinical and public health screening tool for the prevention of CVD deaths.

**Clinical Perspective:** *What is new?:*

- We tested the predictive value of a combination of 113 potential predictors, including 22 clinical blood biomarkers, 20 complete blood counts, 7 anthropometric measures, 51 dietary factors, and 13 cardiovascular health-related questions, beyond traditional risk factors, for CVD mortality in adults in the United States.
- The addition of predictors, specifically blood biomarkers such as glucose, uric acid, bicarbonate, urea nitrogen, total protein, creatinine, calcium, globulin, and phosphorus, improved CVD mortality prediction.

*What are the clinical implications?:*

- Accurate prediction of CVD mortality is essential for identifying those at risk and targeting interventions.
- Our findings highlight the clinical translational utility of predictors, including the biomarkers already well established and routinely applied in clinical practice, for CVD mortality prediction.

## INTRODUCTION

Cardiovascular diseases (CVD) remain major threats to public health in the United States and around the world. Cost-effective primary prevention of CVD relies on the accuracy of risk assessment, specifically, the identification of individuals at high risk of developing CVD. Traditional risk factors, such as age, blood pressure, smoking, serum lipids, and diabetes, as implemented through the Framingham Risk Score and the Pooled Cohort Equations (PCE) for the atherosclerotic CVD risk, are widely used in CVD risk assessment in clinical settings.^1,2^ Approximately half of CVD incidence, however, occurs in people who are not classified in the high-risk group.^3^ The addition of several non-traditional risk factors including ankle-brachial index,^4,5^ C-reactive protein,^6^ and coronary artery calcium score,^7^ have not substantially improved risk stratification. Given that risk estimates are used to guide clinical interventions, it is crucial to improve CVD risk-scoring algorithms to target those at risk for preventive measures.

Clinical blood biomarkers used in the diagnosis and treatment of cardiometabolic diseases, blood counts, anthropometric measures, and lifestyle factors are associated with CVD risk and CVD events. Some individual measures, such as C-reactive protein (CRP) and waist circumference, improve CVD risk prediction.^8–12^ Despite these known associations, no previous studies have developed and validated an integrated risk prediction model using all predictors. Our aim was to quantify the improvement of risk prediction of CVD death, beyond traditional risk factors, through the addition of a combination of 113 potential predictors, including 22 clinical blood biomarkers, 20 complete blood counts, 7 anthropometric measures, 51 dietary factors, and 13 cardiovascular health-related questions, using data from a nationally representative population, the United States National Health and Nutrition Examination Survey (NHANES) in the 2001 to 2016 survey cycles linked with the National Death Index mortality data through 2019.

## METHODS

### Study population

Participants included were from NHANES, which is an ongoing, large, cross-sectional survey to assess the health and nutritional status of the civilian, noninstitutionalized U.S. population. All data and materials are publicly available on the National Center for Health Statistics website (https://www.cdc.gov/nchs/nhanes/index.htm). The protocols for NHANES were approved by the National Center for Health Statistics of the Centers for Disease Control and Prevention Institutional Review Board, and informed consent was obtained from all participants.

Data from eight continuous NHANES cycles (2001-2002, 2003-2004, 2005-2006, 2007-2008, 2009-2010, 2011-2012, 2013-2014, and 2015-2016) were used to develop and validate the risk prediction models based on the data availability. For this analysis, we restricted to 29,181 adults aged 40 years and above because participants younger than 40 have very low cardiovascular risks.^13^ Of these adults, 29,130 were linked to the National Death Index mortality data by the National Center for Health Statistics. We further excluded 7,148 participants with missing information on a set of potential predictors, leaving a total of 21,982 participants for the current analysis. We randomly split the data by a ratio of 8:2 into the training set (N=17,638) for model construction and optimization, and the testing set (N=4,344) for evaluating model performance. We did this random split five times and evaluated model performance each time, with similar results. An overview of the study design is shown in **Figure S1**.

### Outcomes

The vital status and cause of death were determined using the public-use NHANES linked mortality data through December 31, 2019, which utilized probabilistic matching algorithms to link NHANES records with death certificates from the National Death Index. Details of matching criteria and calibration are provided by the National Center for Health Statistics.^14^ Death from cardiovascular causes was determined based on the following *International Classification of Disease-10 codes* (ICD-10): I00–I09, I11, I13, I20–I51, and I60– I69.

### Predictors

Traditional CVD risk factors included age (years), sex (male/female), race/ethnicity (non -Hispanic White, non-Hispanic Black, Hispanic including Mexican American and other Hispanic, and other), current smoking status (yes/no), systolic blood pressure (SBP, mm Hg), serum total cholesterol (mg/dL) and high-density lipoprotein (HDL) cholesterol (mg/dL) concentrations, use of antihypertensive medications (yes/no), and diabetes status (yes/no).^1^ We additionally included predictors from clinical blood biomarkers used in diagnosing and treating cardiometabolic diseases, complete blood counts, anthropometric measures, dietary nutrient intake, and cardiovascular health-related questionnaires. Fasting blood samples were collected in the morning and anthropometric measurements were taken at the mobile examination center. Information on 24-h dietary recall and cardiovascular health was obtained using a computer-assisted data collection and coding system before the physical examination. A detailed description of the laboratory methods and questionnaires is available at the National Center for Health Statistics website (https://www.cdc.gov/nchs/nhanes/index.htm). <u>We further selected</u> predictors based on the variable continuity across multiple survey cycles and availability within each survey cycle to obtain a large sample that can allow development and validation of the prediction models, yielding a total of 122 predictors (9 traditional CVD risk factors and 113 potential predictors) available across all eight survey cycles from 2001-2016. A full list of predictors is shown in **Table 1**.

**Table 1.** A list of predictors in the present study.

|  |
| --- |
| **Traditional cardiovascular disease predictors (n=9)** |
| Age, sex, race/ethnicity, current smoking status, systolic blood pressure, serum total cholesterol, serum high-density lipoprotein (HDL) cholesterol, use of antihypertensive medications, and diabetes. |
| **New cardiovascular disease predictors under investigation in the present study** |
| **Clinical blood biomarkers (n=22)** |
| Albumin, alanine aminotransferase (ALT), aspartate aminotransferase (AST), alkaline phosphatase, bicarbonate, urea nitrogen, calcium, gammaglutamyl transaminase (GGT), glucose, iron, lactate dehydrogenase (LDH), phosphorus, total bilirubin, total protein, triglyceride, uric acid, creatinine, sodium, potassium, chloride, osmolality, and globulin. |
| **Complete blood counts (n=20)** |
| White blood cell count, lymphocyte percent, monocyte percent, segmented neutrophils percent, eosinophils percent, basophils percent, lymphocyte number, monocyte number, segmented neutrophils number, eosinophils number, basophils number, red cell count, hemoglobin, hematocrit, mean cell volume, mean cell hemoglobin, mean cell hemoglobin concentration, red cell distribution width, platelet count, and mean platelet volume. |
| **Anthropometric measures (n=7)** |
| Weight, height, body mass index (BMI), waist circumference, upper leg length, upper arm length, and arm circumference. |
| **Total nutrient intake (n=51)** |
| Energy, protein, carbohydrate, dietary fiber, total fat, total saturated fatty acids, total monounsaturated fatty acids, total polyunsaturated fatty acids, cholesterol, Vitamin A, alpha-carotene, beta-carotene, Vitamin B1, Vitamin B2, niacin, Vitamin B6, total folate, Vitamin B12, Vitamin C, calcium, phosphorus, magnesium, iron, zinc, copper, sodium, potassium, selenium, caffeine, theobromine, alcohol, moisture, saturated fatty acids (SFA) 4:0, SFA 6:0, SFA 8:0, SFA 10:0, SFA 14:0, SFA 18:0, monounsaturated fatty acid (MFA) 16:1, MFA 18:1, MFA 18:1, MFA 20:1, MFA 22:1, polyunsaturated fatty acid (PFA) 18:2, PFA 18:3, PFA 18:4, PFA 20:4, PFA 20:5, PFA 22:5, and PFA 22:6. |
| **Cardiovascular health-related questions (n=13)** |
| Pain or discomfort in chest, pain in chest when walking uphill or in a hurry, pain in chest during an ordinary pace on level ground, severe pain in chest more than half hour, pain in right arm, pain in right chest, pain in neck, pain in upper sternum, pain in lower sternum, pain in left chest, pain in left arm, pain in epigastric area, and shortness of breath on stairs/inclines. |

### Statistical Analysis

We described the distribution of covariates in our sample using mean and standard deviation for continuous covariates. We used number and frequency to describe the distribution of categorical covariates. We calculated bivariate descriptive statistics for the sample that experienced a CVD death relative to the sample that alive or experienced a non-CVD death. We compared the distributions in the overall sample to the training set and testing set.

Participants contributed survival time from the NHANES examination to the date of death for those who died from CVD and participants who died from other causes were right-censored at the date of death, and those who were alive were right-censored at the last follow-up date (December 31, 2019). We implemented three statistical/machine learning algorithms for survival analysis, including Cox proportional hazards regression, elastic-net (ENET) penalized Cox regression,^15^ and random survival forest,^16^ to build prediction models for CVD mortality. Cox proportional hazards regression is the most used model for analyzing the association between predictors and survival time. This conventional method, however, may not perform well in the presence of multicollinearity due to potentially high dimensional predictors or when predictors are correlated.^17^ ENET penalized Cox regression is a sparse penalized Cox regression providing satisfactory performance in handling high dimensional predictors.^15^ ENET shrinks coefficients of “unimportant” predictors toward exact zeroes and thus promises to be a useful tool for variable selection and data dimension reduction. Random survival forest is an ensemble model that grows multiple random decision trees using the bootstrap aggregation (bagging) strategy.^16^ Each tree is grown by recursive splitting of data into smaller subgroups (nodes) that minimize the difference in the cumulative hazard within the group while maximizing the difference between groups. The cumulative hazard and survival probability of a participant are calculated at the terminal node. The global survival probability is predicted by averaging over all trees in a forest.

We built 7 different models for each algorithm in the training set. The first model (Model 1) only included 9 traditional CVD risk factors. We then examined whether the addition of other potential predictors improved the predictive performance by incorporating panels of predictors of clinical blood biomarkers (Model 2), complete blood counts (Model 3), anthropometric measures (Model 4), dietary nutrient intake (Model 5), and cardiovascular health questions (Model 6), separately, on top of Model 1. Finally, Model 7 incorporated all 113 potential predictors from different panels to Model 1 (122 predictors in total). All continuous variables were standardized by subtracting the means divided by the standard deviations, and all categorical variables were coded as dummy variables. For ENET penalized Cox regression, the regularization hyperparameters (λ1 and λ2) were ascertained based on a 20-fold cross-validation for maximum C-index. For random survival forest, hyperparameters including the number of variables to possibly split at in each node, the minimum node size required to attempt a split, and the number of trees in the forest were determined by minimizing prediction error in the out-of-bag data. We additionally calculated variable importance by calculating the difference in out-of-bag prediction error between the model, including a specific predictor, and the model substituting such predictor with a random permutation. A higher variable importance indicates that the variable has a higher predictive ability. The R packages “glmnet”^18^ was used to implement the ENET penalized Cox regression and “randomForestSRC”^19^ was used to implement the random survival forest. Complex survey design of NHANES was not considered due to difficulty in handling of survey weights in these machine learning mechanisms.

In the testing set, we evaluated the performance of each prediction model by calculating Harrell C-index,^20^ to quantify the concordance in predicted and observed survival times between participants. C-index ranges from 0 to 1, and a higher value indicate better risk discrimination of the model. We also assessed the improvement of inclusion of additional predictors in the model in risk reclassification by calculating the net reclassification improvement (NRI).^21^ In this study, the 10 year risk of CVD death was predicted, and NRI was quantified by the proportions of CVD death cases correctly assigned a higher predictive probability of at least 5% and those alive correctly assigned a lower probability of at least 5%.

In the sensitivity analysis, we additionally included serum CRP in the prediction model in addition to the clinical blood biomarkers as one of the most studied inflammatory biomarkers in association with CVD.^22,23^ This analysis was conducted in a subpopulation of NHANES 2001-2010 in which CRP data was available. We also assessed whether adding the quadratic terms and pairwise interactions of clinical blood biomarkers further improved the predictive performance in the Cox proportional hazards regression and ENET penalized Cox regression. All analyses were conducted using R, version 4.0.3 (www.R-project.org).

### Data Availability

All data and materials have been made publicly available at the National Center for Health Statistics website (https://www.cdc.gov/nchs/nhanes/index.htm).

## RESULTS

Among these 21,982 participants in the full study sample at baseline, 1,372 died from CVD during the follow-up, with a median follow-up of 9.3 years (maximum follow-up of 19.2 years) (**Table 2**). Participants who died from CVD were more likely to have been male, non-Hispanic White, and at baseline to have been older ages, have had higher SBP, were more likely to have had diabetes, and be users of antihypertensive medications. Participant characteristics were similar between the training and testing sets. Summary statistics of all predictors at baseline can be found in **Table S1**.

**Table 2.** Traditional cardiovascular risk factors of United States National Health and Nutrition Examination Survey participants at study baseline, by cardiovascular disease mortality status at up to 19.2 years of follow up.

| Characteristic | Total population (N = 21982) |  | Training set (N = 17638) |  | Testing set (N = 4344) |  |
| --- | --- | --- | --- | --- | --- | --- |
|  | CVD death<br>(N = 1372) | Non-CVD<br>death or alive*<br>(N=20610) | CVD death<br>(N = 1094) | Non-CVD<br>death or alive<br>(N = 16544) | CVD death<br>(N = 278) | Non-CVD<br>death or alive<br>(N = 4066) |
| Follow-up, median (range), year | 6.6 (0.1, 18.4) | 9.5 (0.1, 19.2) | 6.6 (0.1, 18.4) | 9.5 (0.1, 19.2) | 6.6 (0.1, 17.0) | 9.4 (0.2, 19.0) |
| Age, mean (SD), year | 71.9 (10.5) | 58.5 (12.1) | 71.8 (10.5) | 58.6 (12.1) | 72.4 (10.5) | 58.3 (12.1) |
| No. (%) female | 559 (40.7) | 10469 (50.8) | 438 (40.0) | 8402 (50.8) | 121 (43.5) | 2067 (50.8) |
| No. (%) racial/ethnic groups |  |  |  |  |  |  |
| Non-Hispanic White | 885 (64.5) | 9950 (48.3) | 700 (64.0) | 7954 (48.1) | 185 (66.6) | 1996 (49.1) |
| Non-Hispanic Black | 257 (18.7) | 4037 (19.6) | 200 (18.3) | 3250 (19.6) | 57 (20.5) | 787 (19.4) |
| Hispanic<sup>†</sup> | 190 (13.9) | 5163 (25.1) | 158 (14.4) | 4155 (25.1) | 32 (11.5) | 1008 (24.8) |
| Other race/ethnic group | 40 (2.9) | 1460 (7.1) | 36 (3.3) | 1185 (7.2) | 4 (1.4) | 275 (6.8) |
| No. (%) current smoker | 256 (18.7) | 3903 (18.9) | 198 (18.1) | 3149 (19.0) | 58 (20.9) | 754 (18.5) |
| SBP, mean (SD), mm Hg | 137.5 (23.3) | 127.7 (19.1) | 137.1 (23.3) | 127.7 (19.1) | 139.4 (23.4) | 127.6 (19.2) |
| Total cholesterol, mean (SD), mg/dL | 194.5 (46.3) | 201.3 (42.1) | 193.1 (45.1) | 201.5 (42.2) | 199.7 (50.9) | 200.7 (41.6) |
| HDL cholesterol, mean (SD), mg/dL | 52.4 (15.8) | 53.7 (16.6) | 52.5 (16.0) | 53.7 (16.7) | 51.7 (15.3) | 53.5 (16.2) |
| No. (%) diabetes | 374 (27.3) | 3345 (16.2) | 294 (26.9) | 2665 (16.1) | 80 (28.8) | 680 (16.7) |
| No. (%) use of antihypertensive medications | 853 (62.2) | 8170 (39.6) | 668 (61.1) | 6533 (39.5) | 185 (66.6) | 1637 (40.3) |

Abbreviation: CVD, cardiovascular disease; SBP, systolic blood pressure; HDL, high-density lipoprotein.
\* Non-CVD death included both alive and death of other causes.
<sup>†</sup> Combined from Mexican American and other Hispanic in NHANES.

### Risk discrimination performance

C-indexes for models only include traditional CVD risk factors (Model 1) were 0.850 for Cox proportional hazards regression, 0.851 for ENET penalized Cox regression, and 0.844 for random survival forest (**Table 3**). The addition of clinical blood biomarkers to the traditional CVD risk factors (Model 2) improved C-index to 0.867 for Cox proportional hazards regression, 0.867 for ENET penalized Cox regression, and 0.853 for random survival forest. Increases in C-indexes were also observed when predictors of blood counts were additionally incorporated (Model 3), that C-indexes were 0.861 for Cox proportional hazards regression, 0.860 for ENET penalized Cox regression, and 0.852 for random survival forest. The inclusion of anthropometric measures, dietary nutrient intake, or cardiovascular health-related question predictors did not improve C-indexes. C-indexes were highest in models including all predictors (Model 7) for Cox proportional hazards regression (0.871) and ENET penalized Cox regression (0.871) but not for random survival forest.

**Table 3.** Prediction models performance in cardiovascular disease mortality risk discrimination by C-index and risk reclassification by net reclassification improvement (NRI). Model performance was assessed in the United States National Health and Nutrition Examination Survey testing set (n=4344).

| Models | Algorithms | C-index | NRI |
| --- | --- | --- | --- |
| Model 1: Traditional<sup>*</sup> | Cox proportional hazards regression | 0.850 | -- |
|  | ENET penalized Cox regression | 0.851 | -- |
|  | Random survival forest | 0.844 | -- |
| Model 2: Traditional + clinical blood measures<sup>†</sup> | Cox proportional hazards regression | 0.867 | 0.132 |
|  | ENET penalized Cox regression | 0.867 | 0.111 |
|  | Random survival forest | 0.853 | 0.101 |
| Model 3: Traditional + complete blood count<sup>‡</sup> | Cox proportional hazards regression | 0.861 | 0.037 |
|  | ENET penalized Cox regression | 0.860 | 0.043 |
|  | Random survival forest | 0.852 | 0.041 |
| Model 4: Traditional + anthropometric<sup>§</sup> | Cox proportional hazards regression | 0.852 | 0.012 |
|  | ENET penalized Cox regression | 0.851 | 0.053 |
|  | Random survival forest | 0.848 | -0.018 |
| Model 5: Traditional + nutrient intake | Cox proportional hazards regression | 0.852 | 0.048 |
|  | ENET penalized Cox regression | 0.853 | 0.022 |
|  | Random survival forest | 0.839 | -0.090 |
| Model 6: Traditional + questionnaire<sup>#</sup> | Cox proportional hazards regression | 0.851 | 0.015 |
|  | ENET penalized Cox regression | 0.850 | 0.035 |
|  | Random survival forest | 0.846 | -0.020 |
| Model 7: All predictors<sup>**</sup> | Cox proportional hazards regression | 0.871 | 0.122 |
|  | ENET penalized Cox regression | 0.871 | 0.107 |
|  | Random survival forest | 0.851 | 0.035 |

Abbreviation: ENET: elastic net; NRI: net reclassification improvement.
<sup>\*</sup> Model 1 included 9 traditional cardiovascular disease risk factors.
<sup>†</sup> Model 2 included all predictors from Model 1 + 22 clinical blood biomarker predictors.
<sup>‡</sup> Model 3 included all predictors from Model 1 + 20 complete blood count predictors.
<sup>§</sup> Model 4 included all predictors from Model 1 + 7 anthropometric predictors.
<sup>||</sup> Model 5 included all predictors from Model 1 + 51 total nutrient intake predictors.
<sup>#</sup> Model 6 included all predictors from Model 1 + 13 cardiovascular health-related question predictors.
<sup>\*\*</sup> Model 7 include all predictors from Model 1 + all 113 predictors.

### Risk reclassification performance

The addition of clinical blood biomarkers to the traditional CVD risk factors (Model 2) improved risk reclassification with an NRI of 0.132 for Cox proportional hazards regression, 0.111 for ENET penalized Cox regression, and 0.101 for random survival forest (**Table 3**). Additional inclusion of blood count predictors improved risk reclassification to a minor degree. No improvement was observed when anthropometric measures, dietary nutrient intake, or cardiovascular health-related question predictors were included in the models. When all predictors were included in the prediction models (Model 7), an improvement in risk reclassification was observed; however, the increase was smaller than when only clinical blood biomarkers were included (Model 2). Hazard ratios associated with a one standard deviation increase in predictors in Cox proportional hazards regression Model 2 are shown in **Table S2**.

### Important predictors

Figure 1. shows the top 20 of all predictors (Model 7) using the random survival forest. Two traditional CVD risk factors-age and SBP were shown on the list, while age was the most important predictor among all predictors. Ten clinical blood biomarkers (bicarbonate, urea nitrogen, total protein, creatinine, calcium, globulin, phosphorus, glucose, osmolality, and bilirubin) were among the top 20 predictors. In addition to clinical blood biomarkers that were prominently featured, four blood count variables (monocyte percent, basophil number, mean platelet volume, lymphocyte number), three dietary nutrient intake variables (MFA 22:1, MFA 20:1, magnesium), and the anthropometric measure of arm circumference were also among the top 20 predictors.

**Figure 1.**
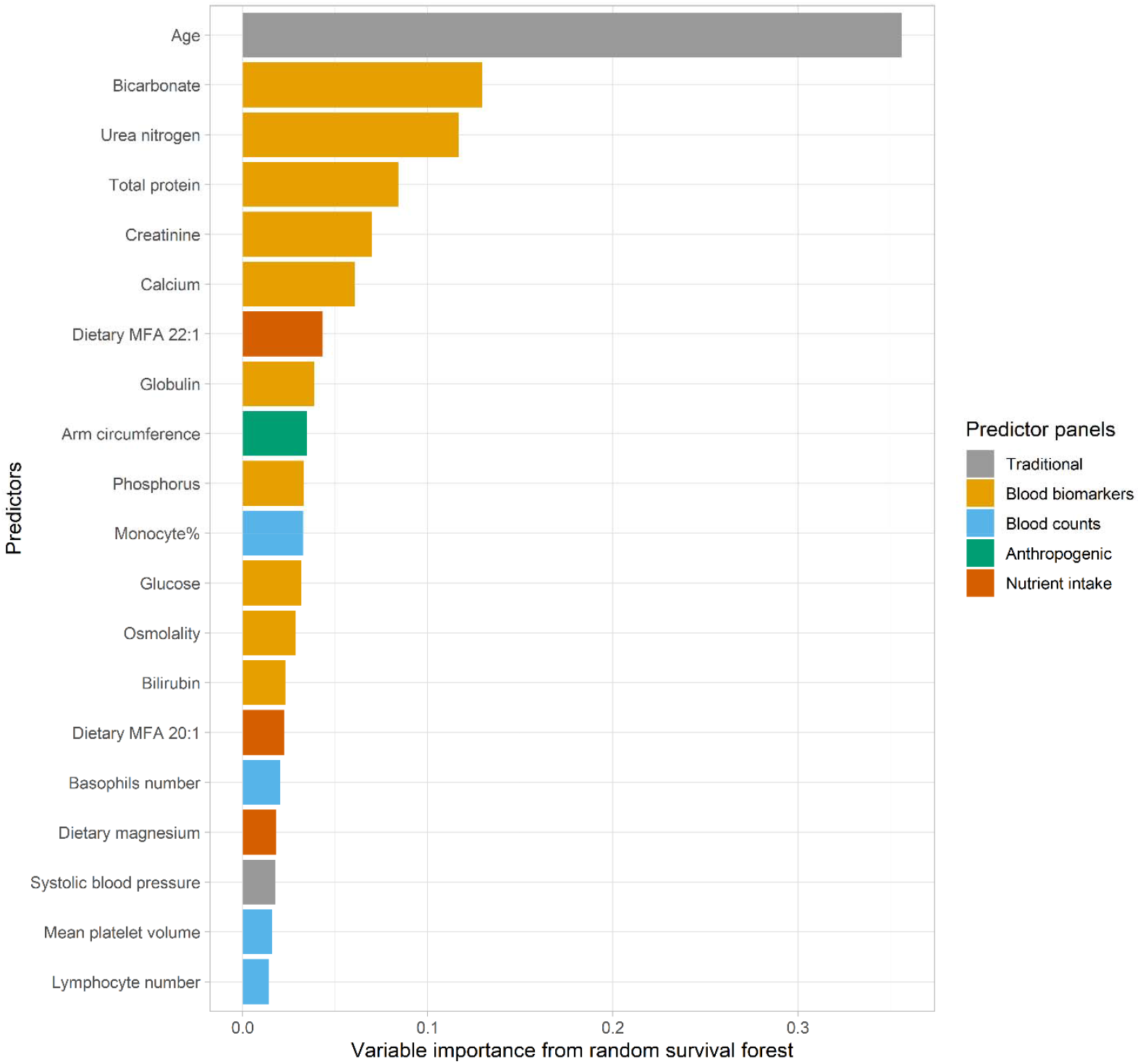
Among all predictors of cardiovascular disease mortality in the United States National Health and Nutrition Examination Survey, the variable importance of the top 20 predictors prioritized from random survival forest analyses.

### Sensitivity analysis

In a subpopulation of 13,620 participants (N=10,926 for the training set and N=2,694 for the testing set) where CRP data was available, including CRP to clinical blood biomarkers slightly improved predictive performance (**Table S3**). In the sensitivity analysis of adding 22 quadratic terms and 231 pairwise interactions of clinical blood biomarkers to the linear terms, no significant improvement in model performance was observed (**Table S4**).

## DISCUSSION

In this large, diverse sample of 21982 U.S. adults aged 40 years and above with up to 19.2 years of longitudinal follow up, the incorporation of a combination of predictors of clinical blood biomarkers, complete blood counts, anthropometric measures, dietary factors, and cardiovascular health-related questions to the model with traditional CVD risk factors improved predictive performance in terms of risk discrimination and reclassification for CVD death. Among all these predictors, clinical blood biomarkers were featured most prominently in predictive performance increases observed in all Cox proportional hazards regression, ENET penalized Cox regression, and random survival forest models.

Clinical blood biomarkers included in the current analysis, such as glucose, CRP, bicarbonate, urea nitrogen, total protein, creatinine, calcium, globulin, and phosphorus are routinely evaluated markers of cardiometabolic, liver, and kidney abnormalities in clinical settings.^24–27^ Associations between these blood biomarkers and CVD-related morbidity and mortality risk have been examined over the last few decades.^24,27,28^ Nevertheless, the predictive value of some of these commonly measured biomarkers has been examined only in a few studies. Blood urea^29^, total protein^30^, and globulin^31^ predict all-cause mortality with moderate predictive performance in CVD patients; however, whether these biomarkers provided additional value to the traditional CVD risk factors has never been evaluated. A study of 2,936 diabetic patients reported that the addition of serum bilirubin to traditional CVD risk factors for predicting CVD mortality increased C-index from 0.713 to 0.729 and showed an 8.6% improvement in net reclassification.^32^ In a cohort of 1,520 consecutive dialysis patients, the addition of serum potassium, calcium, phosphorus, and alkaline phosphatase to traditional CVD risk factors improved C-index from 0.696 to 0.716 for predicting CVD incidence.^33^ More studies have evaluated the predictive value of CRP and serum creatinine. A meta-analysis of 52 cohort studies reported an average of 0.004 increase in C-index and 1.5% NRI for predicting CVD incidence when CRP was added to the traditional CVD risk factors.^25^ Another meta-analysis of 14 studies from the CKD Prognosis Consortium showed an average increase of 0.007 in C-index when serum creatinine was added in the perdition model of CVD mortality using traditional CVD risk factors.^34^ In this study, the inclusion of multiple clinical blood biomarkers improved risk discrimination based on traditional CVD risk factors, as evidenced by around 1.6% increase in the C-index, which is in line with the rationale of using multiple biomarkers involved in multiple disease pathways to improve the risk prediction of CVD death.^23^ Using the information on these routinely measured biomarkers to clinical risk factors may also benefit in identifying subpopulations at higher risks, especially for those classified as “low risk” by clinically used traditional risk models, as evidenced by more than 10% of the study participants were reclassified the predicted risk of more than 5% in the correct direction.

Incorporating complete blood count predictors into the model with traditional CVD risk factors provided a slightly better prediction in terms of risk discrimination and reclassification. This is the first study to assess the predictive value of blood counts for CVD incidence or mortality. Inflammatory blood count markers such as white blood cell count, monocyte percent, and platelet to lymphocyte ratio were associated with poor prognoses in CVD patients.^35^ Mean platelet volume, a marker of platelets’ ability to create thrombus, was also associated with the incidence of CVD.^35^

In our study, adding predictors of anthropometric measures, dietary nutrient intake, and cardiovascular health-related questions to the traditional CVD risk factors, separately, did not add to the risk discrimination performance, though small improvements in risk reclassification were observed. In a meta-analysis of 58 cohorts, the addition of BMI, waist circumference, or waist-to-hip ratio to the risk prediction models for CVD incidence containing traditional risk factors did not improve either risk discrimination or risk reclassification.^12^ In a study combining data from two large cohorts, the prediction model containing information on age, smoking, body mass index, exercise, alcohol, and a composite diet score showed a C-index of 0.72 for predicting CVD incidence; however, whether the composite dietary score itself improved predictive performance was not evaluated.^10^ A recent study of 1,028 patients with non-acute chest pain showed high performance of model incorporating traditional CVD predictors and chest pain-related predictors.^36^ At the same time, it should be acknowledged that a more comprehensive chest pain characteristics, including aspect, localization, radiation, onset, duration, frequency, progress, provoking and relieving factors, and attendant symptoms, were considered in that study which provided more information than NHANES. To note, the small improvement in CVD mortality risk prediction by adding anthropometric measures, dietary nutrient intake, and cardiovascular health-related question-based predictors in our study does not diminish the importance of these variables in CVD morbidity and mortality prevention, as obesity and unhealthy diets are among the major modifiable determinants of CVD.

In our study, three algorithms for survival analysis--Cox proportional hazards regression, ENET penalized Cox regression, and random survival forest models were utilized to build the prediction models. Machine learning algorithms, including ENET penalized Cox regression and random survival forest, were designed to handle the complex correlations between predictors in high-dimensional data and capture the complex predictors-outcome relationships. Nevertheless, in this analysis, ENET penalized Cox regression models showed similar predictive performance as Cox proportional hazards regression models, while random survival forest models did not perform better than Cox proportional hazards regression models. This could reflect that the relationships between predictors considered in our study and CVD mortality may not be complicated enough; thus, the advantages of machine learning algorithms over Cox proportional hazards regression did not play a significant role. Several studies showed that machine learning algorithms better predicted CVD risk over the Cox proportional hazards regression.^37,38^ At the same time, it should be noted that more deep phenotyping predictors such as magnetic resonance imaging markers and electrocardiographic variables were included in these studies. Our findings highlight that there is no universal “best” approach for prediction and that the appropriate modeling strategy is dependent on the different research settings.

Strengths of the present study include using a large and diverse sample of the general U.S. adults, up to 19.2 years of longitudinal follow up, a large number of predictors from different panels, and three different statistical/machine learning algorithms to build prediction models for CVD mortality. We acknowledge that other biomarkers not tested in NHANES such as lipoprotein-associated phospholipase A2 and natriuretic peptides may provide additional information.^39^ Other potential predictors not measured in every NHANES cycle or only available in different subpopulations such as fibrinogen,^25^ physical activity,^40^ and specific environmental toxicants^41^ may also improve the risk prediction. We selected 122 predictors in this analysis and considered data availability in a large sample that can allow the development and validation of the prediction models. Additionally, risk of incident CVD was not predicted due to the cross-sectional nature of NHANES. Our findings encourage confirmations in other longitudinal cohort studies where well-defined longitudinal cardiovascular events are available.

## CONCLUSIONS

Accurate prediction of CVD mortality is essential for identifying those at risk and targeting interventions. Relative to prediction with traditional factors, we observed additionally including multiple predictors with clinical blood biomarkers, complete blood counts, anthropometric measures, dietary factors, and cardiovascular health-related questions improved risk prediction performance for CVD mortality. Specifically, 22 blood biomarkers such as glucose, uric acid, bicarbonate, urea nitrogen, total protein, creatinine, calcium, globulin, and phosphorus contributed the most to improving CVD mortality prediction. Future studies are needed to confirm and validate these findings. The collection of information on these predictors, including the biomarkers, is already well established, and routinely applied in clinical practice, highlighting the clinical translational utility of these predictors for CVD mortality prediction.

## Non-standard Abbreviations and Acronyms

CRP: C-reactive protein
CVD: Cardiovascular diseases
ENET: elastic-net
HDL: high-density lipoprotein
NHANES: National Health and Nutrition Examination Survey
NRI: net reclassification improvement
PCE: Pooled Cohort Equations
SBP: systolic blood pressure

