## Supplemental Material for "Improving the Prediction of Death from Cardiovascular Causes with Multiple Risk Markers"

**Table S1.** Characteristics of United States National Health and Nutrition Examination Survey participants at study baseline, by cardiovascular disease mortality status at up to 19.2 years of follow up.

**Table S2.** Hazard ratios (95% confidence intervals) for a standard deviation increase in predictors in Cox proportional hazards regression Model 2 in the United States National Health and Nutrition Examination Survey testing set (n=4344).

**Table S3.** Cardiovascular disease mortality risk prediction model performance by additional inclusion of C-reactive protein (CRP) to blood biomarkers. Model performance was assessed in the United States National Health and Nutrition Examination Survey testing set (n=4344).

**Table S4.** Cardiovascular disease risk prediction model performance by additional inclusion of quadratic terms and pairwise interactions of blood biomarkers. Model performance was assessed in the United States National Health and Nutrition Examination Survey testing set (n=4344).

**Figure S1**. Flow chart of the study design. NHANES: the National Health and Nutrition Examination Survey; NDI: National Death Index.

**Table S1.** Characteristics of United States National Health and Nutrition Examination Survey participants at study baseline, by cardiovascular disease mortality status at up to 19.2 years of follow up.

| **Characteristics** | **CVD death**  **(N = 1372)** | **Non-CVD death***  **(N=20610)** |
| --- | --- | --- |
| **Clinical blood biomarkers, mean (SD)** |  |  |
| Albumin, g/dL | 4.1 (0.3) | 4.2 (0.3) |
| ALT, IU/L | 22.2 (16.5) | 25.5 (24.2) |
| AST, IU/L | 25.9 (14.2) | 26.3 (19.4) |
| Alkaline phosphotase, IU/L | 75.9 (27.8) | 70.9 (25.0) |
| Bicarbonate, mmol/L | 25.1 (2.5) | 25.1 (2.3) |
| Urea nitrogen, mg/dL | 18.4 (8.8) | 14.3 (5.9) |
| Calcium, mg/dL | 9.4 (0.4) | 9.4 (0.4) |
| GGT, U/L | 33.9 (46.9) | 31.4 (47.5) |
| Glucose, mg/dL | 114.9 (50.1) | 106.1 (40.8) |
| Iron, ug/dL | 82.0 (31.9) | 84.6 (33.9) |
| LDH, U/L | 142.2 (32) | 133.1 (29.9) |
| Phosphorus, mg/dL | 3.7 (0.6) | 3.7 (0.5) |
| Bilirubin, total, mg/dL | 0.8 (0.3) | 0.7 (0.3) |
| Protein, total, g/dL | 7.2 (0.6) | 7.2 (0.5) |
| Triglycerides, mg/dL | 154.6 (98.6) | 159.7 (119.0) |
| Uric acid, mg/dL | 6.1 (1.7) | 5.5 (1.4) |
| Serum creatinine, mg/dL | 1.1 (0.7) | 0.9 (0.4) |
| Sodium, mmol/L | 139.1 (2.8) | 139.2 (2.4) |
| Potassium, mmol/L | 4.1 (0.4) | 4.0 (0.4) |
| Chloride, mmol/L | 102.9 (3.6) | 103.6 (3.0) |
| Osmolatity, mmol/kg | 280.6 (6.3) | 278.9 (5.2) |
| Globulin, g/dL | 3.0 (0.5) | 2.9 (0.5) |
| **Complete blood counts, mean (SD)** |  |  |
| White blood cell count, 1000 cells/uL | 7.3 (2.2) | 7.1 (2.7) |
| Lymphocyte percent, % | 26.8 (8.9) | 30.3 (8.7) |
| Monocyte percent, % | 8.6 (3) | 8.0 (2.3) |
| Segmented neutrophils percent, % | 60.9 (9.8) | 58.1 (9.5) |
| Eosinophils percent, % | 3.1 (2.2) | 2.9 (2.1) |
| Bosophils percent, % | 0.7 (0.5) | 0.7 (0.5) |
| Lymphocyte number, 1000 cells/uL | 1.9 (1) | 2.1 (1.6) |
| Monocyte number, 1000 cells/uL | 0.6 (0.2) | 0.6 (0.2) |
| Segmented neutrophils number, 1000 cells/uL | 4.5 (1.7) | 4.2 (1.7) |
| Eosinophils number, 1000 cells/uL | 0.2 (0.2) | 0.2 (0.2) |
| Basophils number, 1000 cells/uL | 0.04 (0.06) | 0.04 (0.07) |
| Red cell count, million cells/uL | 4.5 (0.5) | 4.6 (0.5) |
| Hemoglobin, g/dL | 14.0 (1.6) | 14.1 (1.5) |
| Hematocrit, % | 41.3 (4.7) | 41.7 (4.2) |
| Mean cell volume, fL | 91.3 (5.8) | 89.9 (5.8) |
| Mean cell hemoglobin, pg | 30.9 (2.3) | 30.5 (2.4) |
| Mean cell hemoglobin concentration, g/dL | 33.8 (0.9) | 33.9 (1) |
| Red cell distribution width, % | 13.4 (1.5) | 13.2 (1.3) |
| Platelet count, 1000 cells/uL | 238.6 (71.6) | 248.9 (67.8) |
| Mean platelet volume, fL | 8.1 (1) | 8.2 (0.9) |
| **Anthropometric measures, mean (SD)** |  |  |
| Weight, kg | 79.6 (20.1) | 81.4 (19.4) |
| Standing Height, cm | 166.5 (10.1) | 166.9 (10.1) |
| Body Mass Index, kg/m^2^ | 28.6 (6.0) | 29.2 (6.1) |
| Waist Circumference, cm | 102.5 (15.0) | 100.7 (14.8) |
| Upper Leg Length, cm | 38.2 (3.9) | 38.3 (4.0) |
| Upper Arm Length, cm | 37.8 (2.7) | 37.4 (2.8) |
| Arm Circumference, cm | 32.0 (5.0) | 33.1 (4.7) |
| **Total nutrient intake, mean (SD)** |  |  |
| Energy, kcal | 1798 (889) | 2013 (914) |
| Protein, gm | 69.8 (35.9) | 78 (39.7) |
| Carbohydrate, gm | 219.0 (109.0) | 243.2 (115.6) |
| Dietary fiber, gm | 14.6 (8.7) | 16.7 (10.3) |
| Total fat, gm | 68.9 (42.7) | 76.8 (44) |
| Total saturated fatty acids, gm | 22.6 (14.8) | 24.6 (15.6) |
| Total monounsaturated fatty acids, gm | 25.2 (16.7) | 27.9 (17) |
| Total polyunsaturated fatty acids, gm | 14.7 (10.8) | 17.3 (11.6) |
| Cholesterol, mg | 268.3 (220.0) | 287.5 (236.5) |
| Vitamin A, RAE, mcg | 593.9 (513.0) | 616.9 (774.4) |
| Alpha-carotene, mcg | 364.3 (975.6) | 410.8 (1171.6) |
| Beta-carotene, mcg | 1982.4 (3342.9) | 2252.9 (4288.5) |
| Thiamin, Vitamin B1, mg | 1.5 (0.8) | 1.5 (0.8) |
| Riboflavin, Vitamin B2, mg | 2.0 (1.2) | 2.1 (1.2) |
| Niacin, mg | 20.8 (12.5) | 23.4 (13.1) |
| Vitamin B6, mg | 1.7 (1.2) | 1.9 (1.2) |
| Total Folate, mcg | 358.0 (219.0) | 387.1 (229.5) |
| Vitamin B12, mcg | 4.8 (5.3) | 5 (7.5) |
| Vitamin C, mg | 80.3 (75.3) | 85.1 (92.3) |
| Calcium, mg | 774.7 (495.5) | 860 (537.9) |
| Phosphorus, mg | 1141.4 (557.0) | 1284 (625) |
| Magnesium, mg | 253.6 (130.3) | 288.9 (142.2) |
| Iron, mg | 13.8 (7.6) | 14.5 (8.3) |
| Zinc, mg | 10.6 (9.4) | 11.1 (8.6) |
| Copper, mg | 1.1 (0.8) | 1.3 (1.3) |
| Sodium, mg | 2927.2 (1535.4) | 3273 (1720) |
| Potassium, mg | 2459.9 (1125.8) | 2645.6 (1232.8) |
| Selenium, mcg | 95.8 (53.2) | 107.1 (61.8) |
| Caffeine, mg | 156.4 (205.1) | 173.2 (219.1) |
| Theobromine, mg | 34.2 (83.0) | 34.7 (77.1) |
| Alcohol, gm | 7.4 (24.9) | 9.4 (26.4) |
| Moisture, gm | 2151.8 (1175.3) | 2653.5 (1420.2) |
| SFA 4:0, gm | 0.5 (0.5) | 0.5 (0.5) |
| SFA 6:0, gm | 0.3 (0.3) | 0.3 (0.3) |
| SFA 8:0, gm | 0.2 (0.2) | 0.2 (0.3) |
| SFA 10:0, gm | 0.4 (0.4) | 0.4 (0.4) |
| SFA 12:0, gm | 0.6 (0.9) | 0.7 (1.2) |
| SFA 14:0, gm | 1.8 (1.6) | 2 (1.7) |
| SFA 16:0, gm | 12.3 (7.8) | 13.4 (8.2) |
| SFA 18:0, gm | 5.9 (3.9) | 6.2 (4) |
| MFA 16:1, gm | 1.0 (0.8) | 1.1 (0.9) |
| MFA 18:1, gm | 23.5 (15.8) | 26 (16) |
| MFA 20:1, gm | 0.2 (0.3) | 0.3 (0.3) |
| MFA 22:1, gm | 0 (0.2) | 0 (0.1) |
| PFA 18:2, gm | 12.9 (9.8) | 15.2 (10.4) |
| PFA 18:3, gm | 1.3 (1.0) | 1.6 (1.2) |
| PFA 18:4, gm | 0.01 (0.03) | 0.01 (0.04) |
| PFA 20:4, gm | 0.1 (0.1) | 0.1 (0.1) |
| PFA 20:5, gm | 0.04 (0.13) | 0.04 (0.13) |
| PFA 22:5, gm | 0.02 (0.04) | 0.02 (0.05) |
| PFA 22:6, gm | 0.1 (0.2) | 0.1 (0.2) |
| **Cardiovascular health-related questions, N (%)** |  |  |
| Pain or discomfort in chest | 463 (33.7) | 5512 (26.7) |
| Pain in chest when walking uphill or in a hurry | 144 (10.5) | 1476 (7.2) |
| Pain in chest during an ordinary pace on level ground | 52 (3.8) | 518 (2.5) |
| Severe pain in chest more than half hour | 131 (9.5) | 1358 (6.6) |
| Pain in right arm | 9 (0.7) | 75 (0.4) |
| Pain in right chest | 14 (1) | 183 (0.9) |
| Pain in neck | 17 (1.2) | 120 (0.6) |
| Pain in upper sternum | 70 (5.1) | 622 (3) |
| Pain in lower sternum | 20 (1.5) | 163 (0.8) |
| Pain in left chest | 43 (3.1) | 372 (1.8) |
| Pain in left arm | 8 (0.6) | 99 (0.5) |
| Pain in epigastric area | 2 (0.1) | 27 (0.1) |
| Shortness of breath on stairs/inclines | 650 (47.4) | 6834 (33.2) |

* Non-CVD death included both alive and death of other causes.

**Table S2.** Hazard ratios (95% confidence intervals) for a standard deviation increase in predictors in Cox proportional hazards regression Model 2 in the United States National Health and Nutrition Examination Survey testing set (n=4344).

| **Clinical blood biomarkers*** | **HR (95% CI)**^†^ |
| --- | --- |
| Albumin, g/dL | 0.61 (0.29, 1.28) |
| ALT, IU/L | 0.79 (0.66, 0.95) |
| AST, IU/L | 1.14 (1, 1.29) |
| Alkaline phosphotase, IU/L | 1.07 (1.02, 1.12) |
| Bicarbonate, mmol/L | 0.93 (0.87, 1.01) |
| Urea nitrogen, mg/dL | 1.05 (0.78, 1.42) |
| Calcium, mg/dL | 0.96 (0.9, 1.03) |
| GGT, U/L | 1.04 (1, 1.08) |
| Glucose, mg/dL | 1.06 (0.78, 1.45) |
| Iron, ug/dL | 0.92 (0.85, 0.99) |
| LDH, U/L | 1.09 (1.05, 1.13) |
| Phosphorus, mg/dL | 1.05 (0.98, 1.12) |
| Bilirubin, total, mg/dL | 1.06 (0.99, 1.13) |
| Protein, total, g/dL | 1.68 (0.52, 5.36) |
| Triglycerides, mg/dL | 0.98 (0.9, 1.07) |
| Uric acid, mg/dL | 1.12 (1.05, 1.2) |
| Serum creatinine, mg/dL | 1.06 (1.02, 1.1) |
| Sodium, mmol/L | 0.94 (0.52, 1.7) |
| Potassium, mmol/L | 1.05 (0.99, 1.12) |
| Chloride, mmol/L | 0.85 (0.77, 0.94) |
| Osmolatity, mmol/kg | 1.13 (0.56, 2.28) |
| Globulin, g/dL | 0.63 (0.2, 2) |

Abbreviation: HR, hazard ratio; CI, confidence interval.

* All clinical blood biomarkers were standardized.

^†^ The model included 9 traditional cardiovascular disease risk factors and 22 clinical blood biomarker predictors.

**Table S3.** Cardiovascular disease mortality risk prediction model performance by additional inclusion of C-reactive protein (CRP) to blood biomarkers. Model performance was assessed in the United States National Health and Nutrition Examination Survey testing set (n=4344).

| **Models** | **Algorithms** | **C-index** | **NRI** |
| --- | --- | --- | --- |
| Model 1* | Cox proportional hazards regression | 0.857 | -- |
|  | ENET penalized Cox regression | 0.855 | -- |
|  | Random survival forest | 0.848 | -- |
| Model 2^†^ | Cox proportional hazards regression | 0.868 | 0.106 |
|  | ENET penalized Cox regression | 0.864 | 0.060 |
|  | Random survival forest | 0.851 | 0.151 |
| Model 2 + CRP | Cox proportional hazards regression | 0.869 | 0.106 |
|  | ENET penalized Cox regression | 0.868 | 0.078 |
|  | Random survival forest | 0.852 | 0.142 |

Abbreviation: ENET: elastic net; NRI: net reclassification improvement; CRP: C-reactive protein.

*Model 1 include 9 traditional cardiovascular disease risk factors.

^†^ Model 2 include all predictors from Model 1 + 22 clinical blood biomarker predictors.

**Table S4.** Cardiovascular disease risk prediction model performance by additional inclusion of quadratic terms and pairwise interactions of blood biomarkers. Model performance was assessed in the United States National Health and Nutrition Examination Survey testing set (n=4344).

| **Models** | **Algorithms** | **C-index** | **NRI** |
| --- | --- | --- | --- |
| Model 1* | Cox proportional hazards regression | 0.850 | -- |
|  | ENET penalized Cox regression | 0.851 | -- |
| Model 2^†^ | Cox proportional hazards regression | 0.867 | 0.132 |
|  | ENET penalized Cox regression | 0.867 | 0.111 |
| Model 2 + 22 quadratic terms + 231 pairwise interactions of blood biomarkers | Cox proportional hazards regression | 0.851 | 0.123 |
|  | ENET penalized Cox regression | 0.867 | 0.085 |

Abbreviation: ENET: elastic net; NRI: net reclassification improvement.

*Model 1 include 9 traditional cardiovascular disease risk factors.

^†^ Model 2 include all predictors from Model 1 + 22 clinical blood biomarker predictors.

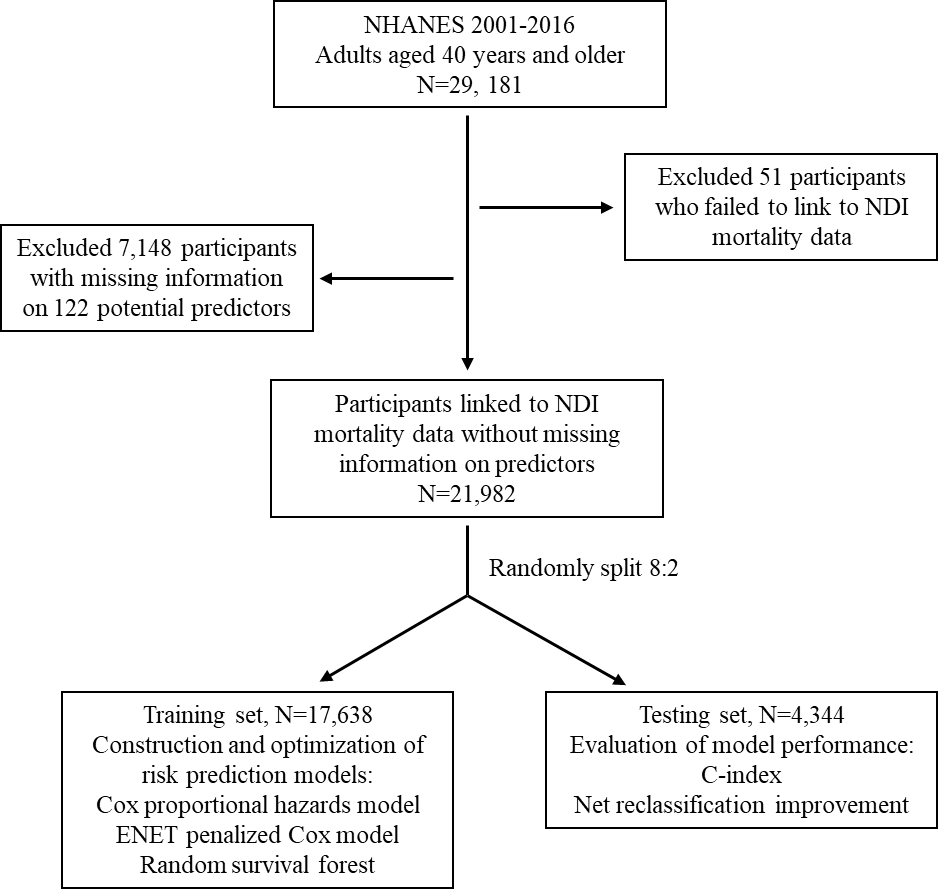

**Figure S1**. Flow chart of the study design. NHANES: the National Health and Nutrition Examination Survey; NDI: National Death Index.
